# Immune modulating drug MP1032 with SARS-CoV-2 antiviral activity in vitro: A potential multi-target approach for prevention and early intervention treatment of COVID-19

**DOI:** 10.1101/2020.11.03.20216580

**Authors:** Sara Schumann, Astrid Kaiser, Ferdinando Nicoletti, Katia Mangano, Paolo Fagone, Eduard van Wijk, Yu Yan, Petra Schulz, Beate Ludescher, Michael Niedermaier, Joerg von Wegerer, Pia Rauch, Christian Setz, Ulrich Schubert, Wolfgang Brysch

## Abstract

At least since March 2020, the multiorgan disease COVID-19 has a firm grip on the world. Although most of the cases are mild, patients from risk populations could develop a cytokine storm, which is characterized by a systemic inflammatory response leading to acute respiratory distress syndrome and organ failure. The present paper will introduce the small molecule MP1032, describe its mode of action, and give rationale why it is a promising option for prevention/treatment of SARS-CoV-2-induced cytokine storm. MP1032 is a phase-pure anhydrous polymorph of 5-amino-2,3-dihydro-1,4-pthalazinedione sodium salt that exhibits good stability and bioavailability. The physiological action of MP1032 is based on a multi-target mechanism including localized, self-limiting antioxidant activities that were demonstrated in a model of lipopolysaccharide (LPS)-induced joint inflammation. Furthermore, immune-regulatory and PARP-1 modulating properties, coupled with antiviral effects against SARS-CoV-2 were shown in various cell models. Efficacy has been preclinically elucidated in LPS-induced endotoxemia, a model with excessively activated immune responses that shares many similarities to COVID-19. So far, during oral clinical development with three-months daily administrations, no serious adverse drug reactions occurred highlighting the outstanding safety profile of MP1032.

## 1. Introduction

The multiorgan disease COVID-19 (Corona virus disease 2019) is caused by infection with SARS-CoV-2. It may result in the acute respiratory distress syndrome (ARDS) and multiple organ failure that have been ascribed to a cytokine storm (also called cytokine release syndrome or macrophage overactivation syndrome), a systemic inflammatory response associated with an overstimulated immune system [1,2]. However, not all people exposed to SARS-CoV-2 are infected and not all infected patients develop severe COVID-19. Among 1099 patients analyzed in Wuhan, 16% progress to the severe phase showing respiratory failure that requires mechanical ventilation [3].

One of the biggest unanswered questions is why some people develop severe disease, whilst others do not. More and more data support the notion that there is a skewed mortality towards elderly men with underlying diseases [3,4]. Elderly persons often show a diminished/impaired pulmonary immunity with inadequate innate and adaptive immune responses. The so called inflammaging is characterized by elevated reactive oxygen species (ROS), inflammatory cytokines and acute phase reactants (e.g. C-reactive protein) that lead to continuous low-grade inflammation accompanied by an overall decline in innate immune responses [5]. The process of inflammaging together with the presence of other age-related diseases, such as diabetes and cardiovascular diseases, is thought to be associated with increased severity and mortality of viral infections in elderly people [6].

SARS-CoV-2 infects epithelial cells of the upper respiratory tract via the receptor for angiotensin-converting enzyme 2 (ACE2) triggering innate response mechanisms [7]; at later stages, the virus may disseminate to the lower respiratory tract [8]. Upon introduction of viral proteins and RNA into host cells, endoplasmic reticulum stress and mitochondrial dysfunction are induced promoting ROS production [9]. Extensive ROS lead to oxidative stress and increase cytokine production (e.g. IL-6) facilitating the dreaded cytokine storm in severe COVID-19 cases that is a major factor for high mortality, multiorgan failure, ARDS, and disseminated intravascular coagulation [10]. On systemic level, high neutrophil to lymphocyte ratios are reported in critically ill COVID-19 patients that have been found to predict in-hospital mortality [11]. It is hypothesized that this neutrophilia also excessively generates ROS (oxidative burst) that exacerbate the host immunopathological response resulting in a more severe disease [12].

Back to the cellular level, maintenance of normal homeostasis and counteraction of detrimental ROS effects relies among others on effective DNA repair. Several DNA repair mechanisms exist, one is the Poly (ADP-ribose) polymerase 1 (PARP1) that is responsible for ADP-ribosylation. PARP-1 is activated by binding to irregular/damaged DNA [13] that is often the result of extensive ROS production under inflammatory conditions [14]. Hence, PARP enzymes are involved in a wide variety of cellular processes like cell death, immune function, antiviral response, and autophagy [15]. In regard to SARS-CoV-2 infection, it is interesting that PARP-1 inhibition has been shown to limit inflammation-induced tissue damage, including acute lung injury in animal models [16] and PARP-1 inhibitors are therefore discussed as treatment option for COVID-19 [15].

Similar to SARS-CoV and MERS-CoV, there is currently no clinically proven specific antiviral agent available for SARS-CoV-2 infection and the medical need for safe, relatively cheap and easily distributable drugs is high. Our data suggest that the small molecule MP1032 exerts antiviral activity against SARS-CoV-2. MP1032 is a phase-pure anhydrous polymorph of 5-amino-2,3-dihydro-1,4-pthalazinedione sodium salt [17] that is very stable, water soluble and has good bioavailability via different administration routes. It is currently under development at MetrioPharm (Zurich, Switzerland) reaching clinical Phase II for the treatment of psoriasis showing an outstanding safety profile. The physiological action of MP1032 is based on a multi-target mechanism that depends on localized, self-limiting antioxidant activities and PARP-1 modulating properties. Efficacy has been shown in preclinical models with excessively activated immune responses (such as lipopolysaccharide (LPS)-induced endotoxemia) predisposing MP1032 for the treatment/prevention of SARS-CoV-2-induced cytokine storm. The present paper will introduce this small molecule, describe its mode of action and give rationale why it is a promising treatment option for COVID-19.

## 2. Results

### 2.1 MP1032, a self-regulating, highly specific ROS scavenger with PARP-1 inhibitor activity

Based on its chemical properties, MP1032 is thought to scavenge excessive inflammation-induced ROS leading to MP1032 oxidation and elevated photon emission. This photon emission enhances the normally occurring signal and is detectable by ultra-weak photon emission (UPE) imaging.

To evaluate this effect in detail, joint inflammation was induced in the right ankle of mice by intra-articular LPS injection; as control, PBS was injected into the left ankle joint. At maximum inflammation, MP1032 was applied systemically via i.p. injection and UPE was monitored. The scavenging reaction was observed between 1 min and 30 min after MP1032 injection, although it was strongest after 15 min. Figure 1 indicates MP1032-mediated photon emission in the right ankle joint, directly at the site of LPS-induced inflammation. In contrast, PBS application into the left ankle joint did not result in inflammation and therefore no UPE was detectable. This unique and self-limiting mechanism is based on the fact that excessive ROS production creates intracellularly a highly localized alkaline environment [18], which deprotonates and thereby activates circulating MP1032 directly at the inflammatory site. After restoring cellular physiologic redox balance via the scavenging of excessive ROS, the environment returns to normal pH, MP1032 action is ceased and the UPE signal declines. This signal decline is primarily due to rapidly decreasing MP1032 activation, rather than tissue drug concentrations, which persist considerably longer as shown by profound pharmacokinetic data (data not shown).

**Figure 1:**
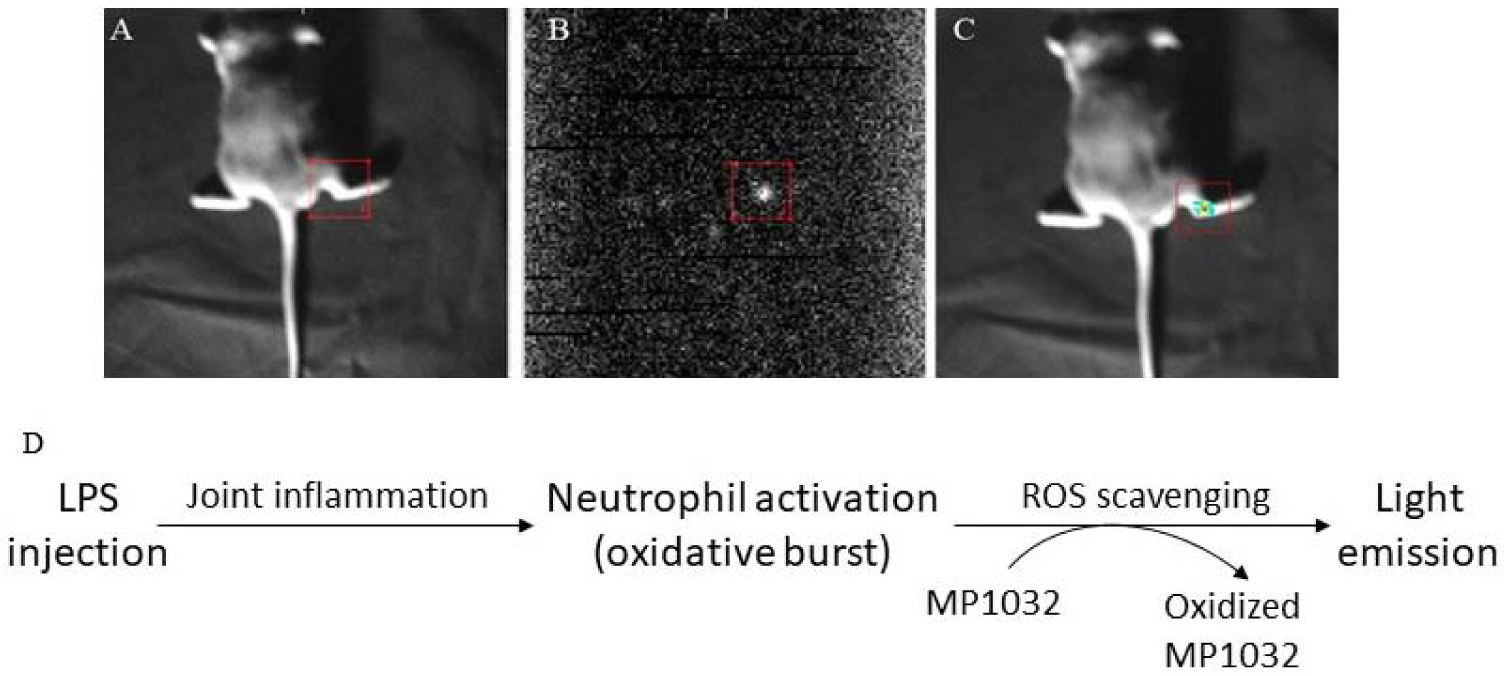
Effects of MP1032 on LPS-induced production of reactive oxygen species (ROS). Using male DBA/1J mice, joint inflammation was induced in the right ankle by intra-articular injection of 10 µg LPS; as control, PBS was injected into the left ankle joint. After two days, 100 mg/kg MP1032 was applied systemically via i.p. injection and ultra-weak photon emission was measured 15 min post-dose. (A) position image, (B) photon image, (C) merged image with pseudo-color, and (D) proposed mechanism of ROS scavenging. Pictures show one representative mouse from a total of 3 mice.

Oxidative stress induces DNA damage and thereby activates repair mechanisms such as PARP-1. We therefore investigated whether MP1032 can affect PARP-1 activity. At first, a coupled enzyme assay with fluorometric read-out was used to determine the effect on human PARP-1 activity in a cell-free system. In principle, PARP activity is indicated by NAD^+^ depletion and reduced fluorescence. Hence, reduced PARP-1 activity due to PARP inhibition rescues NAD^+^ and leads to higher fluorescence values. For MP1032 an inhibitory activity on human PARP-1 was detected in the low micromolar range (IC_50_ = 1.55 μM), while the established PARP inhibitor 4-amino-1,8-naphthalimide (4-ANI) showed an IC_50_ of 0.13 μM (Figure 2A). In a second experiment it was evaluated whether MP1032 is also able to inhibit PARP-1 activity in differentiated HL-60 cells. By measuring the amount of poly-ADP-ribosylated proteins, it was shown that differentiated but untreated HL60 showed already high levels of PARP-1 activity (Figure 2B). Further, it indicates that poly-ADP-ribosylation is not massively increased by LPS stimulation, an effect that was previously described [19]. Nevertheless, upon treatment with 1 mM MP1032 or 50 μM 4-ANI, the amount of poly-ADP-ribosylation proteins decreased dramatically by over 90%.

**Figure 2:**
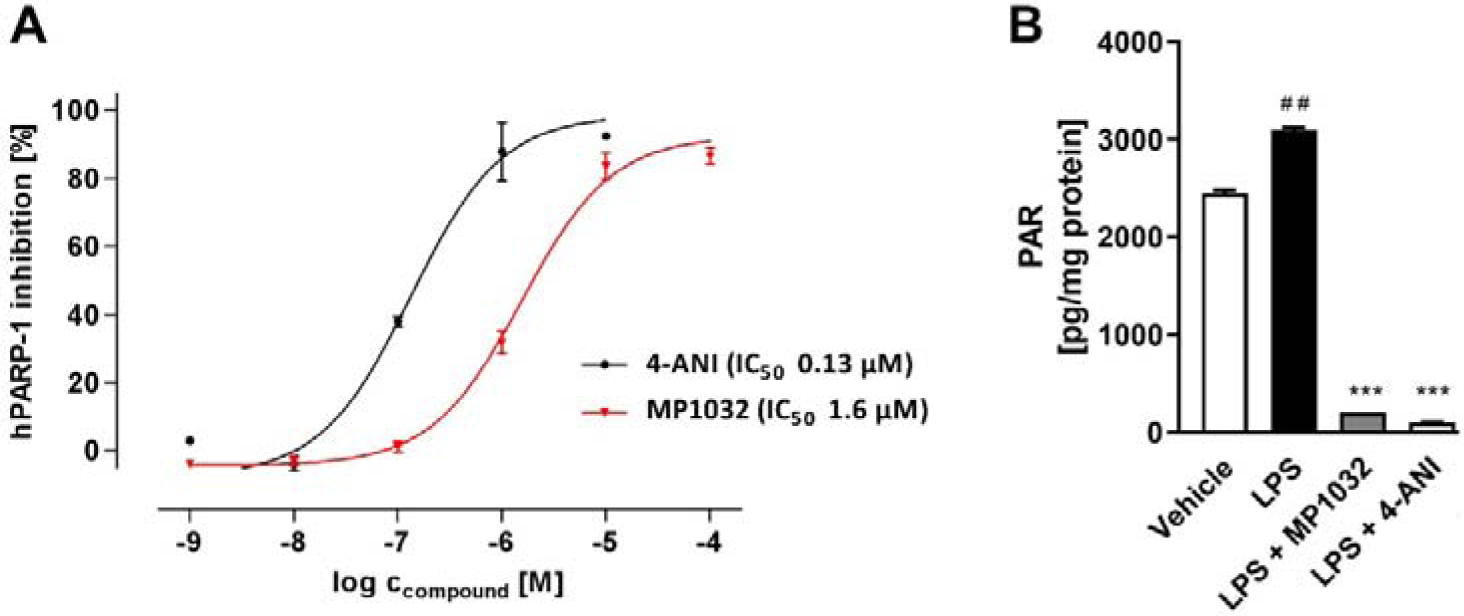
Effects of MP1032 on PARP-1. (A) Cell-free human PARP-1 enzyme assay with MP1032 or 4-amino-1,8-naphthalimide (4-ANI). Measured RFU raw data were normalized by using a NAD^+^ standard curve and expressed as inhibition of hPARP-1 [%]. (B) Poly-ADP-ribosylation (PAR) of proteins in differentiated HL60 cells upon treatment with 1 mM MP1032 or 50 μM 4-ANI. HL60 were differentiated for 24 hours with phorbol 12-myristate 13-acetate and then treated for another 24 hours with MP1032 and 4-ANI under LPS inflammatory conditions. Total cellular protein was isolated and subjected to PAR ELISA. PAR values were normalized to the respective protein content. Data are expressed as mean ± SEM, n= 4/group. Statistical analyses were performed using T-Test; ^##^p<0.01 (Vehicle vs. LPS), ***p<0.001 (treatments vs. LPS).

Overall, these studies demonstrate that MP1032 works as ROS scavenger acting directly and exclusively at the site of inflammation. Furthermore, MP1032 seems to modulate PARP-1 activity directly and independently from its ROS scavenging capacities.

### 2.2 MP1032 attenuates LPS-induced proinflammatory cytokine activation in vitro and in vivo

During bacterial as well SARS-CoV-2-induced inflammation, oxidative stress and PARP-1 activation are tightly linked to the induction of proinflammatory cytokine production. The potential effect of MP1032’s ROS scavenging and PARP-1-inhibiting properties on the downstream activation of pro-inflammatory cytokines was tested in HL-60 and murine/human primary immune cells.

The human promyelocytic leukemia cell line, HL-60 was differentiated along the macrophage/monocyte linage and LPS stimulation with either 0.1 µg/ml or 1 µg/ml LPS led to a dose-dependent TNFα and IL-6 induction (Figure 3A). Pre-treatment with 1 mM MP1032 reduced the secretion of both cytokines. Using 1 µg/ml LPS, MP1032 reduced the TNFα and IL-6 levels about 39% and 52%, respectively. When 0.1 µg/ml LPS was used, MP1032 led mainly to a decrease of TNFα (about 53%).

**Figure 3:**
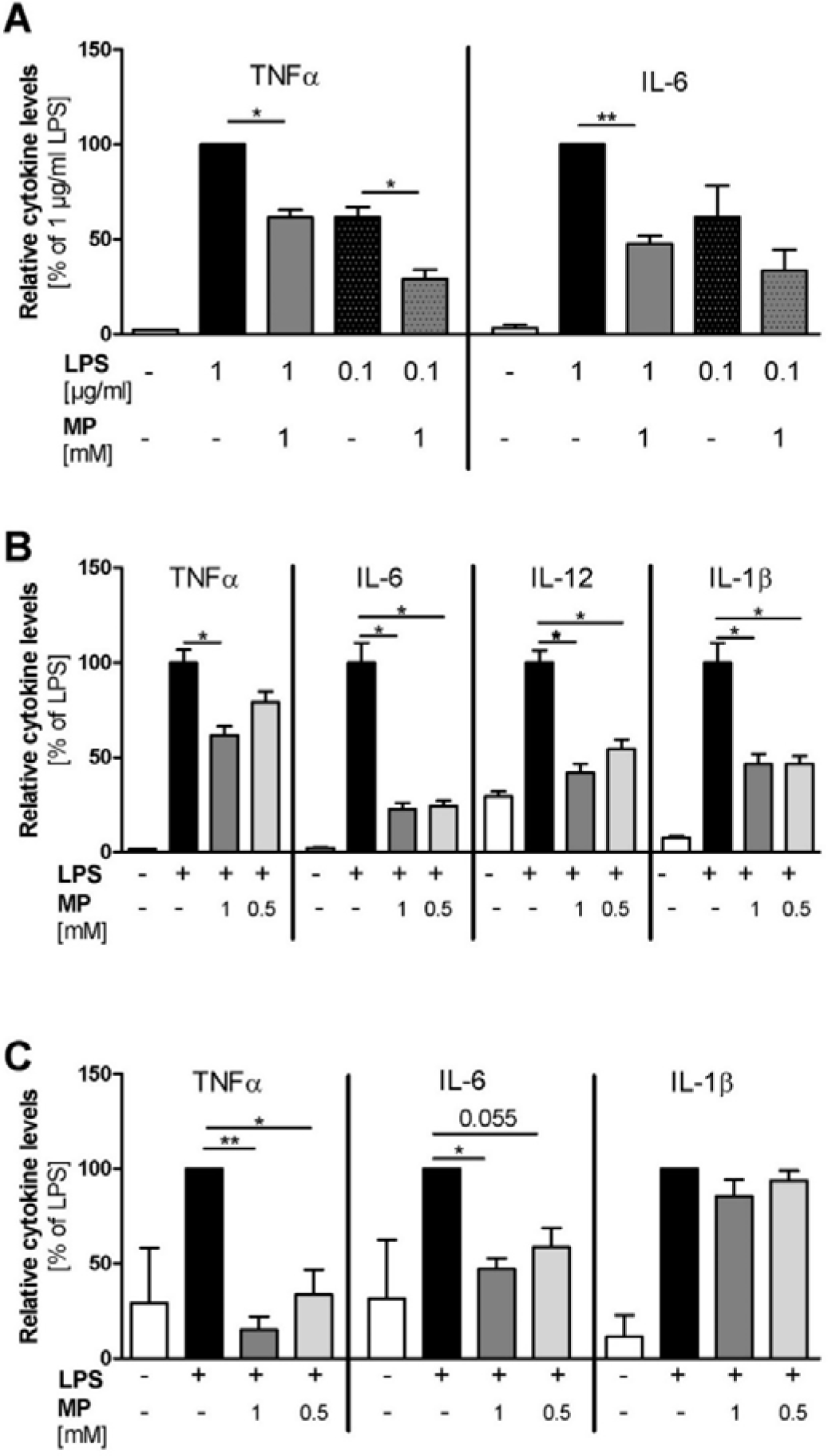
MP1032 attenuates LPS-induced cytokine release in vitro. (A) human HL60 cells were differentiated for 24 h using 50 ng/ml phorbol 12-myristate 13-acetate (PMA). One hour prior to LPS stimulation (with 0.1 µg/ml or 1 µg/ml LPS), differentiated cells were treated with 1 mM MP1032. (B) murine peritoneal macrophages were isolated from female C57Bl/6 mice four days after i.p. injection of 3% thioglycolate medium. One hour prior to LPS stimulation (0.1 µg/ml), isolated cells were pre-treated with 0.5 or 1 mM MP1032. (C) human peripheral blood mononuclear cells were isolated from blood buffy coats by density gradient centrifugation. One hour prior to stimulation with 0.1 µg/ml LPS, cells were treated with 0.5 or 1 mM MP1032. Cell-free supernatants were collected from all cell types after 24 h and secreted cytokine levels were detected by ELISA. Data are expressed as mean + SEM relative to the LPS-treated vehicle group that was normalized to 100%. (A/B) n= 2, (C) n=3. Statistical analyses were performed using unpaired T Test with or without Welch’s correction; *p<0.05, **p<0.01.

Using LPS-stimulated murine peritoneal macrophages, we assessed the effects of MP1032 in a primary cell model. While MP1032 did not affect cytokine secretion of unstimulated murine PMs (data not shown), LPS-induced proinflammatory cytokine concentrations were reduced by MP1032 pre-treatment (Figure 3B). In detail, 1 mM MP1032 reduced levels of TNFα, IL-6, IL-12, and IL1-β by about 40%, 80%, 60%, and 50%, respectively. Interestingly, for all cytokines similar MP1032 effects were observed after 48 h and 72 h of culture. Furthermore, these effects were validated in PMs of a second mouse strain (Balb/c mice) and a clear dose-dependency was observed with six different MP1032 concentrations ranging from 1 mM to 0.25 mM (Supplementary Figure 1).

Using human PBMCs, MP1032 at concentrations of 0.5 mM and 1 mM reduced LPS-induced TNFα and IL-6 secretion in a dose-dependent manner (Figure 3C). IL-1β levels were reduced to a much lower extent, which did not reach statistical significance. Notably, the oxidized form of MP1032 (3-aminophthalic acid sodium salt) did not have any effect on LPS-induced TNFα and IL-6 secretion indicating that the ROS scavenging properties described under 2.1 and an intact structure are mandatory to achieve suppression of proinflammatory cytokine production.

The efficacy of MP1032 to inhibit the secretion of proinflammatory cytokines in vitro prompted us to evaluate its in vivo efficacy. The used sublethal LPS-challenged model represents a model of endotoxic shock that is pathophysiologically characterized by dysregulated secretion of proinflammatory cytokines [20]. As depicted in Figure 4, MP1032 pretreatment significantly reduced plasma cytokine levels compared to vehicle-treated mice by 50% (TNFα) and 25% (IL-6). The positive control dexamethasone was administered two times (instead of once as for MP1032) and reduced TNFα as well as IL-6 concentrations by 70% and 45%, respectively.

**Figure 4:**
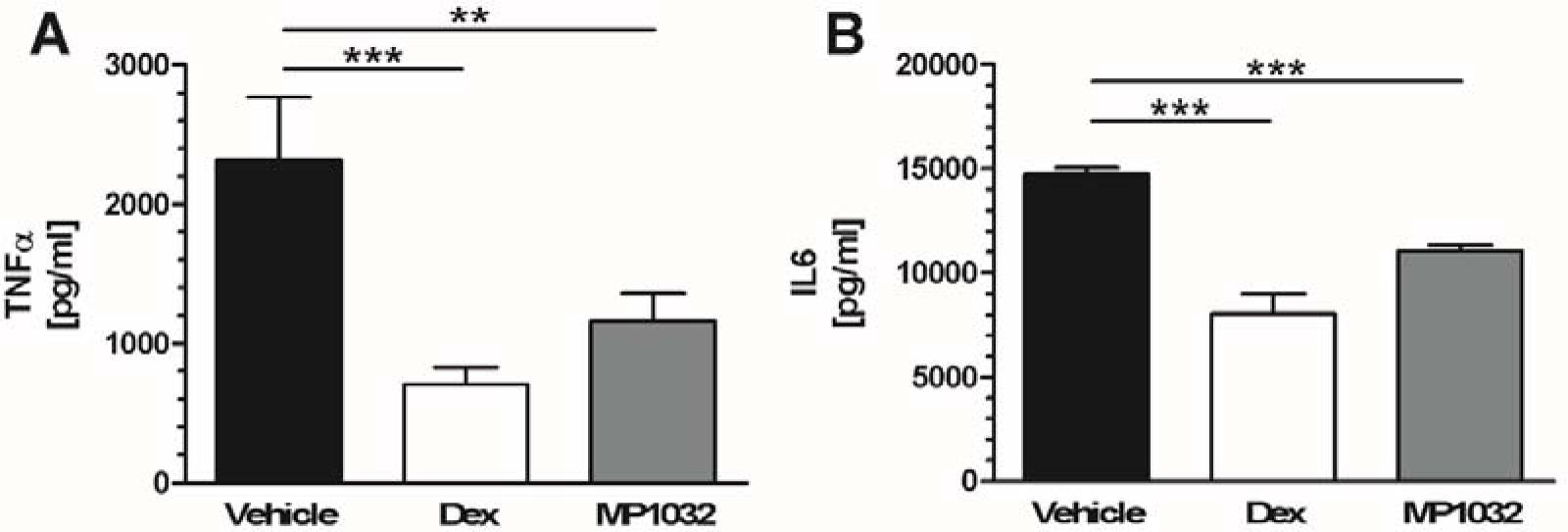
Proinflammatory cytokine levels in LPS-treated mice. Fifteen minutes prior to LPS challenge (300 µg/mouse; i.p.), female CD1 mice were treated via i.p. injection with 0.5 mg/kg MP1032. Dexamethasone (Dex; 0.3 mg/kg) was dosed twice, 24 h and 1 h before LPS application; as vehicle, water for injection was used. Mice were sacrificed 2 h after LPS challenge and plasma TNFα (A) and IL-6 (B) concentrations were determined by ELISA. Data are expressed as mean ± SEM, n=9-10/group. Statistical analyses were performed using Mann Whitney Test; **p<0.01, ***p<0.001.

Taken together, MP1032 reduces proinflammatory cytokine levels in various immune cells and attenuates LPS-induced endotoxemia in vivo.

### 2.3 MP1032 exhibits an antiviral activity against SARS-CoV-2

To analyze if MP1032 possesses antiviral activity against SARS-CoV-2, Vero B4 cells were infected with SARS-CoV-2_PR-1_ and treated with MP1032. Three days post infection, cell culture supernatants were harvested and virus production was analyzed by Western blot. Treatment with MP1032 led to strong reduction of SARS-CoV-2 replication. At 1 mM MP1032, the production of progeny virions was inhibited by about 70% and at 2 mM almost completely blocked (Figure 5A). To control for potential unspecific effects on cell viability, a WST-1 assay was performed in uninfected Vero B4 cells showing that MP1032 concentrations effectively suppressing SARS-CoV-2 replication do not affect cell viability (Figure 5B).

**Figure 5:**
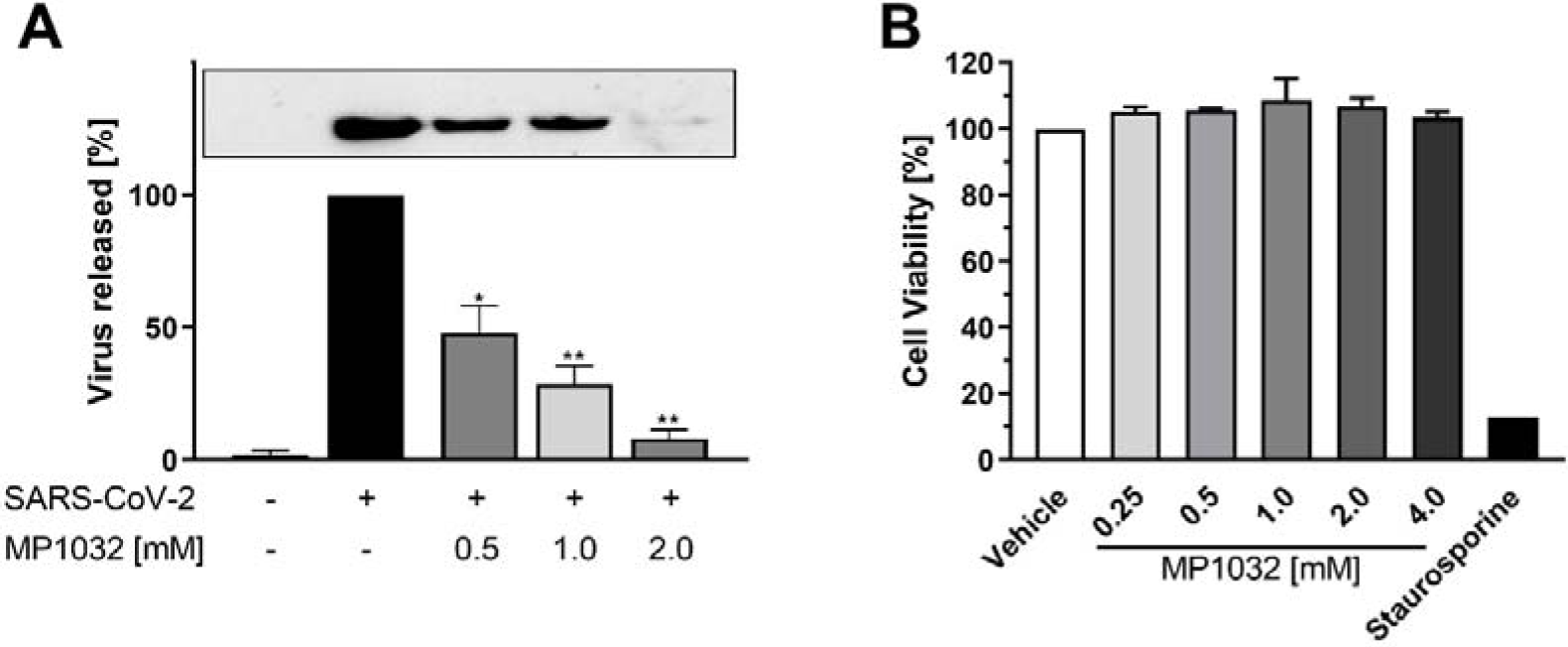
Influence of MP1032 on SARS-CoV-2 replication and viability of Vero B4 cells. Confluent monolayers of Vero B4 cells were infected with a 100-fold dilution of the field isolate SARS-CoV-2_PR-1_ for 1h and afterwards treated with MP1032. (A) 72 hours post infection, SARS-CoV-2_PR-1_ nucleocapsid was determined in supernatants by Western Blot analysis (a representative blot is shown). (B) Viability of uninfected and treated cells was assessed by the water-soluble tetrazolium salt 1 assay; staurosporine (1 µM) served as positive control. Data are expressed as mean ± SEM, n= 3/group. Statistical analyses were performed using unpaired T Test with Welch’s correction; **p<0.01, *p<0.05.

Thus, these cumulative data implicate that MP1032 exhibits strong antiviral activity against SARS-CoV-2, while having no impact on Vero B4 cell viability.

### 2.4 Clinical safety data

The data presented above clearly indicate that MP1032 is a promising treatment option for SARS-CoV-2-infection and the prevention of the COVID-19 stage. To further emphasize this, clinical safety data obtained from a randomized, double-blind, placebo-controlled phase II study in psoriasis patients are presented here. In total, 155 patients were randomized receiving either 300 mg MP1032, 150 mg MP1032, or placebo twice daily over a study period of 12 weeks. Safety evaluation revealed that MP1032 at both doses was safe and generally well-tolerated with no clinically important safety issues being identified. No deaths occurred and only three treatment-emergent serious adverse events (SAEs) occurred in the placebo group. Most of the treatment-emergent adverse events (TEAEs) were ‘unlikely’ or ‘not related’ to MP1032 and patients receiving 300 mg MP1032 showed a statistically significant reduction in the incidence of adverse events compared to patients receiving placebo (Figure 6).

**Figure 6:**
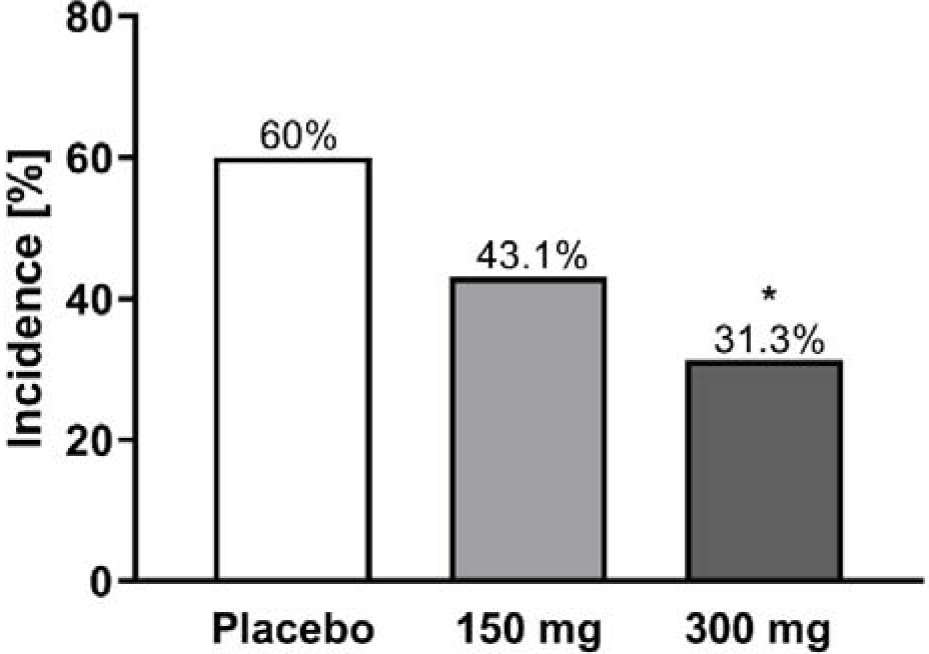
Incidence of treatment-emergent adverse events in a clinical phase II study. In total, 155 patients with moderate to severe chronic plaque psoriasis were randomized receiving either 300 mg MP1032, 150 mg MP1032, or placebo twice daily of a period of 12 weeks (EudratCT-No:2017-003484-36). Adverse events were coded according to the Medical Dictionary for Regulatory Activities. Statistical analyses were performed using two-sided Fisher’s exact test; *p<0.05.

To summarize this, MP1032 was successfully evaluated in the clinical setting showing an exceptional safety profile with good tolerability.

## 3. Discussion

The World Health Organization declared the COVID-19 outbreak, caused by SARS-CoV-2, a pandemic on March 11, 2020. Given this immense health risk, several drugs have been employed ranging from antivirals, antibiotics, biologics, and corticosteroids up to antioxidants [21,22]. Despite these and a massive international initiative to develop SARS-CoV-2 vaccines, there still remains an unabated urgent need for effective pharmacologic interventions that prevent COVID-19 patients from clinical decline towards the need for intensive care and assisted ventilation. As of this writing, no such safe, convenient, and widely applicable treatment meeting these criteria has been identified or approved.

Here we present data on MP1032, a small molecule drug which combines localized, auto-regulated ROS-scavenging and immune-modulating effects with specific antiviral properties against SARS-CoV-2.

To date, 146 subjects received MP1032 during oral clinical development up to phase II (EudraCT-Nos. 2014-004606-15, 2015-005159-28, 2017-003484-36). Besides a demonstrated trend of a dose dependent anti-inflammatory effect in those exploratory psoriasis trials, no serious adverse events occurred in the MP1032 treatments groups highlighting its outstanding safety profile (all reported adverse drug reactions are summarized in Supplementary Table 1).

The profound antioxidant and immune-modulating properties of MP1032 originally prompted development in the field of autoimmune inflammatory diseases (like psoriasis or rheumatoid arthritis) and age-related degenerative diseases caused by chronic low-grade inflammation (inflammaging). Similarly, alveolar inflammation and acute lung injury in COVID-19 are driven by cellular pathologies, particularly with respect to oxidative stress and the pattern of cytokines involved in the cytokine release syndrome [23,24]. Several antibodies directed against these pro-inflammatory cytokines or their respective receptors have already been evaluated in the clinical setting, so far with moderate success only. This may be due to the fact that these biologics suppress the host immune system, which is detrimental in the early non-severe stages where the adaptive immune response helps to eliminate the virus and protects against disease progression to severe stages [25]. The importance of treatment timing with immune suppressants was demonstrated by the RECOVERY trial. In this study, dexamethasone resulted in lower mortality among severe COVID-19 cases who were receiving respiratory support, while in patients who were not receiving invasive mechanical ventilation, dexamethasone treatment was harmful and resulted in a higher mortality rate [26]. In contrast to immune-suppressing biologics or corticosteroids, MP1032 acts as an immune regulator since it does not completely suppress cytokine induction, which serves an important role during anti-infective response. The redox-balancing mechanism of MP1032 rather curtails the overshoot of the immune response. Such a pharmacodynamic profile lends itself for early intervention treatment, where preserving a physiologic immune response and inhibiting an overshoot are essential for preventing disease progression.

Similar to chronic inflammatory diseases, several respiratory viruses induce a dysregulated ROS formation by disrupting antioxidant mechanisms leading to an unbalanced oxidant: antioxidant ratio and subsequent oxidative cell damage [27]. Early use of antioxidants, such as vitamin C and N-acetyl cysteine is therefore discussed as potential option to prevent/treat COVID-19 [28,29] and several studies are currently ongoing (e.g. ClinicalTrials.gov Identifier: NCT04344184, NCT04323514, NCT04374461, NCT04419025). In contrast to these antioxidants, which at high concentrations tend to scavenge even physiologically necessary ROS, MP1032 action is self-regulating and highly localized. This unique mechanism is based on the fact that excessive ROS production creates intracellularly a highly localized alkaline environment [18]. Due to its chemical properties, circulating MP1032 is activated by this pH shift only at the inflammatory site. As MP1032 scavenges excessive ROS, the pH normalizes and MP1032 activation shuts down. Hence, excessive ROS are scavenged, but physiologically necessary ROS levels are spared, ensuring that there is no impairment of cellular signaling.

In vitro evaluation of antiviral activity directed against SARS-CoV-2 is preferably performed in Vero cells [30]. Using this model, we could show that MP1032 suppresses virus replication in a dose-dependent manner while the drug has no effect on cell viability. The molecular basis for this anti-viral effect still needs to be evaluated. It is conceivable that MP1032 inhibits virus entry via acting on ACE2 receptor as it was proposed for N-acetyl cysteine [31]. MP1032 could further attenuate prolonged virus replication by preventing oxidative stress [32] or by limiting ADP-ribosylation of the viral nucleocapsid protein via PARP-1 inhibition [33]. Additional studies should be conducted to evaluate this in more detail.

## 4. Material and Methods

### 4.1 MP1032

MP1032 (5-amino-2,3-dihydro-1,4-pthalazinedione sodium salt) polymorph was synthesized at a specialized manufacturer (ChemCon, Germany) in a multistep procedure. The polymorph was formed in the last reaction step and its polymorphic purity was confirmed by X-ray powder diffraction as previously described [17].

### 4.2 In vitro studies

#### 4.2.1 HL-60 cells

As described by Xu et al. [34], the human leukemic cell line HL-60 was differentiated using 50 ng/ml phorbol 12-myristate 13-acetate (PMA, Sigma-Aldrich) for 24 h. To confirm successful differentiation along the monocytic/macrophage lineage, HL-60 were stained according to the method of Pappenheim. One hour prior to LPS stimulation (with 0.1 µg/ml or 1 µg/ml LPS; Sigma-Aldrich, Germany), differentiated cells were treated with 1 mM MP1032 (or 50 µM 4-ANI, for PARP-1 activity). After 24 h, cell-free supernatants were collected and stored at −20°C for cytokine quantification, while the cells were lysed for PARP-1 assay.

#### 4.2.2 Murine peritoneal macrophages

Female C57Bl/6 mice (6-8 weeks old) were purchased from Envigo (San Petro al Natisone, Udine, Italy) and housed under standard laboratory conditions (specific-pathogen free) at the animal facility of the University of Catania (Catania, Italy). This experiment was performed in accordance with the Directive 2010/63/UE (enforced by the Italian D. Lgs 26/2014); physical facilities and equipment for accommodation and care of animals were in accordance with the provisions of EEC Council Directive 86/609. Food and water were applied *ad libitum*. Animal handling and the study protocol were in accordance with international guidelines and approved by the local Institutional Animal Care and Use Committee.

Four days after i.p. injection of 3% thioglycolate medium (St Louis, MO USA), murine peritoneal macrophages (PMs) were isolated according a published protocol [35]. Cell purity was confirmed by flow cytometry (CD14 expression) and isolated PMs were allowed to adhere overnight. One hour prior to LPS stimulation (0.1 µg/ml; *Escherichia coli* O55:B5, St Louis, MO USA), cells were pre-treated with different MP1032 concentrations (ranging from 1 mM to 0.025 mM). Two independent experiments (each with three mice; cells were pooled) were performed and secreted cytokine levels were determined 24, 48 and 72 h after LPS stimulation using ELISA technique.

#### 4.2.3 Human peripheral blood mononuclear cells (PBMCs)

PBMCs were isolated from buffy coats of three different healthy blood donors (Institute for transfusion medicine, Suhl, Germany) by density gradient centrifugation (Ficoll Hypaque, density 1.077 g/ml, Biochrome). One hour prior to stimulation with 0.1 µg/ml LPS (from *Escherichia coli* O111:B4; Sigma Aldrich, Taufkirchen, Germany), PBMCs were treated with different concentrations of MP1032 (1 mM and 0.5 mM). After LPS stimulation, the PBMCs were incubated for 24 hours. Finally, the supernatants of the PBMC culture were collected and frozen at −20°C until cytokine concentrations were determined by ELISA.

#### 4.2.4 Vero B4 cell culture and infection

Vero B4 cells (National institute of health, USA) were maintained in Dulbecco’s Modified Eagle’s Medium (DMEM) containing 10% (v/v) inactivated fetal calf serum (FCS), 2 mM l-glutamine, 100 U/mL penicillin, and 100 μg/mL streptomycin. Confluent monolayers of Vero B4 cells were infected in FCS-free DMEM with a 100-fold dilution of the field isolate SARS-CoV-2_PR-1_ (isolated from a 61 years old patient six days after the presumed date of infection and two days after start of mild COVID-19 symptoms; see [36]) for 1h and afterwards treated with MP1032. 72 hours post infection, virus-containing cell culture supernatants were harvested and released virions were purified through 20% (w/v) sucrose cushion (20,000× g, 4 °C, 90 min). After washing with PBS, the pellet was dissolved in SDS sample buffer and used for Western Blot analysis. Viability of uninfected and treated cells was assessed by WST-1 assay (Roche) according to the manufacturer’s instructions.

### 4.3 In vivo studies

#### 4.3.1. Intra-articular inflammation and ultra-weak photon emission imaging

The study was performed at the animal facility of the Changchun University of Chinese Medicine. The animals were kept under standard laboratory conditions (water and food *ad libitum*). Animals were single housed according to the Chinese legislation; animal handling and the study protocol were in accordance with international guidelines and approved by the local Institutional Animal Care and Use Committee. Cages were sterilized and filled with wood shavings, cellulose as bedding material. Automatically controlled environmental conditions are set to maintain temperature at 20 – 24°C with a relative humidity of 50 – 60%, 12 hours light-dark cycle.

Male DBA/1J mice aged 6-7 weeks received intra-articular injection of LPS (1×10^−5^ g LPS solubilized in 20 μL PBS) into the right ankle joint through the Achilles tendon using a 30-gauge needle, while an identical volume of PBS was injected into the left ankle joint. After two days, mice received i.p. injection of 2.5 mg MP1032, which corresponds for a final dose of 100 mg/kg b.w.. UPE imaging using a highly sensitive electron-multiplying CCD camera system (iXon 888 EM+; Andor Technology Ltd, Belfast, Northern Ireland) was performed 15 min after MP1032 injection: The mice were anesthetized using a Matrx VMR small animal anaesthesia device (Midmark) via isoflurane vapour (3% during induction and 2% during experiment); the duration of anaesthesia was kept as short as possible (within 35 min). A position image of each mouse was taken and after removing the red-light torch, UPE images were recorded in complete darkness.

#### 4.3.2 Sublethal LPS model

This experiment was performed in accordance with the Directive 2010/63/UE (enforced by the Italian D. Lgs 26/2014) at the animal facility of the University of Catania (Catania, Italy); physical facilities and equipment for accommodation and care of animals were in accordance with the provisions of EEC Council Directive 86/609. The animals were kept under standard laboratory conditions (non-specific pathogen free) in a climate-controlled room (20-24°C, relative air humidity 30-70%) with a 12-h light:dark cycle; food and water was applied *ad libitum*. CD1 mice (female, 6 weeks old) were purchased from Harlan Laboratories (San Pietro al Natisone, Udine, Italy). They were allowed to adapt to the animal facility for at least one week before commencing the study. Fifteen minutes prior to i.p. LPS challenge (300 µg/mouse; from *Salmonella enterica*, Cod L6011, Sigma, Milano, Italy), mice were treated via i.p. injection with 0.5 mg/kg MP1032. As control, dexamethasone (Dex; 0.3 mg/kg, Soldesam, Labotatorio Biologico Milanese, Varese, Italy) was dosed twice, 24 h and 1 h before LPS application. As vehicle, water for injection was used. Mice were sacrificed 2 h after LPS challenge and plasma TNFα and IL-6 concentrations were determined by ELISA.

### 4.4 Clinical phase II trial

In a randomized, double-blind, placebo-controlled study, two oral doses of MP1032 were evaluated over a period of 12 weeks in patients with moderate to severe chronic plaque psoriasis (PASI 10–20). In total, 155 patients were randomized receiving either 300 mg MP1032, 150 mg MP1032, or placebo twice daily (EudratCT-No:2017-003484-36). Adverse events were coded according to the Medical Dictionary for Regulatory Activities.

### 4.5 Cytokine measurement

To measure cytokine concentration in cell culture supernatants, the following ELISA kits were used: For the human HL-60 cells/PMBCs: BioLegend (San Diego, CA, USA), and for murine PMs: LifeSpan BioSciences, Inc. (Seattle, WA, USA). Plasma TNFα and IL-6 were determined using eBioscience or Bender Medsystem (Prodotti Gianni, Milano, Italy). ELISA assays were performed according to the manufacturer’s instructions. Data of the in vitro studies are expressed relative to the LPS-treated vehicle group that was normalized to 100%.

### 4.6. PARP-1 activity in HL-60 cells

PARP-1 activity in human HL-60 cells was determined using the PARP in vivo Pharmacodynamic Assay II (Trevigen, USA) according to the manufacturer’s instructions. Poly-ADP-ribosylation was normalized to the protein content measured in the cell lysates.

### 4.7 Cell-free PARP-1 enzyme assay

Cell-free human PARP-1 enzyme activity was determined in a coupled enzyme assay with fluorometric read-out. In a first step, 200 nM NAD^+^ were mixed with the PARP mix consisting of 1.25 μg activated (partly cleaved) DNA (Sigma#D4522) and 1.0 U/well hPARP-1 (Trevigen #4668-100-01) in assay buffer (50 mM Tris and 2 mM MgCl_2_ (pH 8.0)). Subsequently, MP1032 or 4-ANI prepared in assay buffer were added to the reaction mixture. Each assay was performed with a NAD^+^ standard curve (0 – 100 nM NAD^+^) and a PARP minus control for data analysis. After 90 min, alcohol dehydrogenase/diaphorase mix consisting of 50 mU alcohol dehydrogenase, 5 mU diaphorase, 50 μM resazurin and 2% EtOH (v/v) was added. Fluorescence kinetic (5 min steps for 40 min) was measured using Synergy2 plate reader (BioTek, Germany) with λ_exc_ 540/25 nm, λ_em_ 590/20 nm and 550 nm mirror. Data were calculated using NAD^+^ standard curve and expressed as percent hPARP-1 inhibition.

### 4.8 SDS-PAGE and Western Blotting

Protein samples were separated by SDS-PAGE, transferred onto nitrocellulose membranes, blocked with 3% bovine serum albumin, and incubated with the appropriate primary antibody. The SARS-CoV-2 Nucleocapsid antibody (Genetex, GTX135357) and the anti-rabbit secondary antibody coupled to horseradish peroxidase (Dianova, Germany) were used.

### 4.9 Statistical analyses

All data are presented as mean ± SEM. Statistical calculation was performed using GraphPad Prism (GraphPad Software, La Jolla, USA). Normal distribution was tested using the Kolmogorov-Smirnov test. Unpaired T-Test (with or without Welch’s correction depending on the comparability of variances) or Mann-Whitney Test were used to assess the direct treatment effects vs. the vehicle group. To assess the incidence of treatment-emergent adverse events in the clinical phase II study, two-sided Fisher’s exact test was used. Differences with p<0.05 were considered statistically significant.

## 5. Conclusion

No clinically proven specific antiviral agent is currently available for SARS-CoV-2 infection and thus the medical need for safe and easily distributable drugs is high. Here, we show that MP1032, a phase-pure anhydrous polymorph of 5-amino-2,3-dihydro-1,4-pthalazinedione sodium salt exerts potent immune-modulatory, self-regulated ROS scavenging, and SARS-CoV-2 specific antiviral properties. This pharmacodynamic profile which simultaneously addresses several crucial pathophysiological processes of a SARS-CoV-2 infection renders MP1032 a promising candidate for the treatment and possibly prevention of COVID-19. This assumption should be tested in a clinical trial.

## Data Availability

The data that support the findings of this study are available from the corresponding author (SS).

## 6. Acknowledgments

The authors would like to thank David Kosel for his excellent support in experimental planning, monitoring, and analyses.

## 7. Author contributions

Conceptualization, S.S:, A.K., F.N., P.S., E.vW., J.vW., C.S., U.S. and W.B.; formal analysis, S.S., F.N., K.M., P.F., Y.Y., P.S.,C.S. and U.S.; investigation, F.N. K.M., P.F., P.R., C.S., E.vW. and Y.Y.; writing—original draft preparation, S.S.; writing—review and editing, A.K., F.N., K.M., P.F., E.vW., Y.Y., P.S., B.L., M.N., J.vW., C.S., U.S., W.B.; visualization, S.S.; project administration, S.S., A.K., F.N., P.S., B.L., J.vW., E.vW., Y.Y., C.S., U.S. and M.N; supervision, A.K., P.S.. All authors have read and agreed to the published version of the manuscript.

## 10. Abbreviations

4-ANI: 4-amino-1,8-naphthalimide
ACE2: Angiotensin-converting enzyme 2
ARDS: Acute respiratory distress syndrome
COVID-19: Corona virus disease 2019
CD4: Cluster of differentiation 4
Dex: Dexamethasone
DMEM: Dulbecco’s Modified Eagle’s Medium
ELISA: Enzyme-linked Immunosorbent Assay
EtOH: Ethanol
FCS: Fetal calf serum
IL: Interleukin
LPS: Lipopolysaccharide
PARP-1: Poly (ADP-ribose) polymerase 1
PASI: Psoriasis Area and Severity Index
PBMC: Peripheral blood mononuclear cells
PBS: Phosphate buffered saline
PM: Peritoneal macrophages
PMA: Phorbol 12-myristate 13-acetate
RFU: Relative fluorescence units
ROS: Reactive oxygen species
SAE: Serious adverse event
SARS-CoV-2: Severe acute respiratory syndrome coronavirus type 2
SDS-PAGE: Sodium dodecyl sulfate polyacrylamide gel electrophoresis SEM Standard error of the mean
TEAE: Treatment-emergent adverse event
TNFα: Tumor necrosis factor-alpha
UPE: Ultra-weak photon emission
WST-1: Water-soluble tetrazolium salt 1 assay

## References

1. Huang, C., Wang, Y., Li, X., Ren, L., Zhao, J., Hu, Y., Zhang, L., Fan, G., Xu, J., Gu, X., et al. Clinical features of patients infected with 2019 novel coronavirus in Wuhan, China. The Lancet, 395(10223), 497–506 2020, doi:10.1016/S0140-6736(20)30183-5.

2. Li, X., Geng, M., Peng, Y., Meng, L., Lu, S. Molecular immune pathogenesis and diagnosis of COVID-19. J. Pharm. Anal. 2020, 10, 102–108, doi:10.1016/j.jpha.2020.03.001.

3. Guan, W.-J., Ni, Z.-Y., Hu, Y., Liang, W.-H., Ou, C.-Q., He, J.-X., Liu, L., Shan, H., Lei, C.-L., Hui, D.S.C., et al. Clinical Characteristics of Coronavirus Disease 2019 in China. N. Engl. J. Med. 2020, 382, 1708–1720, doi:10.1056/NEJMoa2002032.

4. Williamson, E.J., Walker, A.J., Bhaskaran, K., Bacon, S., Bates, C., Morton, C.E., Curtis, H.J., Mehrkar, A., Evans, D., Inglesby, P., et al. Factors associated with COVID-19-related death using OpenSAFELY. Nature 2020, 584, 430–436, doi:10.1038/s41586-020-2521-4.

5. Lowery, E.M., Brubaker, A.L., Kuhlmann, E., Kovacs, E.J. The aging lung. Clin. Interv. Aging 2013, 8, 1489–1496, doi:10.2147/CIA.S51152.

6. Nasi, A., McArdle, S., Gaudernack, G., Westman, G., Melief, C., Rockberg, J., Arens, R., Kouretas, D., Sjölin, J., Mangsbo, S. Reactive oxygen species as an initiator of toxic innate immune responses in retort to SARS-CoV-2 in an ageing population, consider N-acetylcysteine as early therapeutic intervention. Toxicol. Rep. 2020, 7, 768–771, doi:10.1016/j.toxrep.2020.06.003.

7. Li, F., Li, W., Farzan, M., Harrison, S.C. Structure of SARS coronavirus spike receptor-binding domain complexed with receptor. Science 2005, 309, 1864–1868, doi:10.1126/science.1116480.

8. Zhou, F., Yu, T., Du, R., Fan, G., Liu, Y., Liu, Z., Xiang, J., Wang, Y., Song, B., Gu, X., et al. Clinical course and risk factors for mortality of adult inpatients with COVID-19 in Wuhan, China: a retrospective cohort study. The Lancet 2020, doi:10.1016/S0140-6736(20)30566-3.

9. Shi, C.-S., Nabar, N.R., Huang, N.-N., Kehrl, J.H. SARS-Coronavirus Open Reading Frame-8b triggers intracellular stress pathways and activates NLRP3 inflammasomes. Cell Death Discov. 2019, 5, 101, doi:10.1038/s41420-019-0181-7.

10. Shi, Y., Wang, Y., Shao, C., Huang, J., Gan, J., Huang, X., Bucci, E., Piacentini, M., Ippolito, G., Melino, G. COVID-19 infection: the perspectives on immune responses. Cell Death Differ. 2020, 27, 1451–1454, doi:10.1038/s41418-020-0530-3.

11. Fu, J., Kong, J., Wang, W., Wu, M., Yao, L., Wang, Z., Jin, J., Wu, D., Yu, X. The clinical implication of dynamic neutrophil to lymphocyte ratio and D-dimer in COVID-19: A retrospective study in Suzhou China. Thromb. Res. 2020, 192, 3–8, doi:10.1016/j.thromres.2020.05.006.

12. Laforge, M., Elbim, C., Frère, C., Hémadi, M., Massaad, C., Nuss, P., Benoliel, J.-J., Becker, C. Tissue damage from neutrophil-induced oxidative stress in COVID-19. Nat. Rev. Immunol. 2020, 20, 515–516, doi:10.1038/s41577-020-0407-1.

13. Bai, P. Biology of Poly(ADP-Ribose) Polymerases: The Factotums of Cell Maintenance. Molecular cell 2015, 58, doi:10.1016/j.molcel.2015.01.034.

14. Péter Bai; László Virág. Role of poly(ADP[ribose) polymerases in the regulation of inflammatory processes. FEBS Letters 2012, 586, 3771–3777, doi:10.1016/j.febslet.2012.09.026.

15. Curtin, N., Bányai, K., Thaventhiran, J., Le Quesne, J., Helyes, Z., Bai, P. Repositioning PARP inhibitors for SARS-CoV-2 infection(COVID-19); a new multi-pronged therapy for acute respiratory distress syndrome? Br. J. Pharmacol. 2020, 177, 3635–3645, doi:10.1111/bph.15137.

16. Pazzaglia, S., Pioli, C. Multifaceted Role of PARP-1 in DNA Repair and Inflammation: Pathological and Therapeutic Implications in Cancer and Non-Cancer Diseases. Cells 2019, 9, doi:10.3390/cells9010041.

17. Martin, T., Greim, D., Milius, W., Niedermaier, M., Ludescher, B., Wegerer, J. von; Brysch, W., Bärwinkel, K., Senker, J., Breu, J. The Same at a First Glance: The Diffractogram of a New Polymorph of Anhydrous Sodium Luminolate Almost Perfectly Resembles the Diffraction Trace of an Already Known Polymorph. Z. anorg. allg. Chem. 2015, 641, 332–338, doi:10.1002/zaac.201400604.

18. Ikebuchi, Y., Masumoto, N., Tasaka, K., Koike, K., Kasahara, K., Miyake, A., Tanizawa, O. Superoxide anion increases intracellular pH, intracellular free calcium, and arachidonate release in human amnion cells. J. Biol. Chem. 1991, 266, 13233–13237.

19. Bhatia, M., Kirkland, J.B., Meckling-Gill, K.A. Modulation of poly(ADP-ribose) polymerase during neutrophilic and monocytic differentiation of promyelocytic (NB4) and myelocytic (HL-60) leukaemia cells. Biochem. J. 1995, 308 (Pt 1), 131–137, doi:10.1042/bj3080131.

20. Nicoletti, F., Mancuso, G., Cusumano, V., Di Marco, R., Zaccone, P., Bendtzen, K., Teti, G. Prevention of endotoxin-induced lethality in neonatal mice by interleukin-13. Eur. J. Immunol. 1997, 27, 1580–1583, doi:10.1002/eji.1830270639.

21. Fajgenbaum, D.C., Khor, J.S., Gorzewski, A., Tamakloe, M.-A., Powers, V., Kakkis, J.J., Repasky, M., Taylor, A., Beschloss, A., Hernandez-Miyares, L., et al. Treatments Administered to the First 9152 Reported Cases of COVID-19: A Systematic Review. Infect. Dis. Ther. 2020, 1–15, doi:10.1007/s40121-020-00303-8.

22. Costela-Ruiz, V.J., Illescas-Montes, R., Puerta-Puerta, J.M., Ruiz, C., Melguizo-Rodríguez, L. SARS-CoV-2 infection: The role of cytokines in COVID-19 disease. Cytokine Growth Factor Rev. 2020, 54, 62–75, doi:10.1016/j.cytogfr.2020.06.001.

23. Georg Schett; Bernhard Manger;David Simon; Roberto Caporali. COVID-19 revisiting inflammatory pathways of arthritis. Nat Rev Rheumatol 2020, 16, 465–470, doi:10.1038/s41584-020-0451-z.

24. Tan, Y., Qi, Q., Lu, C., Niu, X., Bai, Y., Jiang, C., Wang, Y., Zhou, Y., Lu, A., Xiao, C. Cytokine Imbalance as a Common Mechanism in Both Psoriasis and Rheumatoid Arthritis. Mediators Inflamm. 2017, 2017, 2405291, doi:10.1155/2017/2405291.

25. Derek W. Cain; John A. Cidlowski. After 62 years of regulating immunity, dexamethasone meets COVID-19. Nat Rev Immunol, 1–2, doi:10.1038/s41577-020-00421-x.

26. Dexamethasone in Hospitalized Patients with Covid-19 — Preliminary Report. N. Engl. J. Med. 2020, doi:10.1056/NEJMoa2021436.

27. Gjyshi, O., Bottero, V., Veettil, M.V., Dutta, S., Singh, V.V., Chikoti, L., Chandran, B. Kaposi’s sarcoma-associated herpesvirus induces Nrf2 during de novo infection of endothelial cells to create a microenvironment conducive to infection. PLoS Pathog. 2014, 10, e1004460, doi:10.1371/journal.ppat.1004460.

28. Cheng, R.Z. Can early and high intravenous dose of vitamin C prevent and treat coronavirus disease 2019 (COVID-19)? Med. Drug Discov. 2020, 5, 100028, doi:10.1016/j.medidd.2020.100028.

29. Schönrich, G., Raftery, M.J., Samstag, Y. Devilishly radical NETwork in COVID-19: Oxidative stress, neutrophil extracellular traps (NETs), and T cell suppression. Advances in Biological Regulation 2020, 77, 100741, doi:10.1016/j.jbior.2020.100741.

30. Jeon, S., Ko, M., Lee, J., Choi, I., Byun, S.Y., Park, S., Shum, D., Kim, S. Identification of Antiviral Drug Candidates against SARS-CoV-2 from FDA-Approved Drugs. Antimicrob. Agents Chemother. 2020, 64, doi:10.1128/AAC.00819-20.

31. Flora, S. de; Balansky, R., La Maestra, S. Rationale for the use of N-acetylcysteine in both prevention and adjuvant therapy of COVID-19. FASEB J. 2020, doi:10.1096/fj.202001807.

32. Khomich, O.A., Kochetkov, S.N., Bartosch, B., Ivanov, A.V. Redox Biology of Respiratory Viral Infections. Viruses 2018, 10, doi:10.3390/v10080392.

33. Grunewald, M.E., Fehr, A.R., Athmer, J., Perlman, S. The coronavirus nucleocapsid protein is ADP-ribosylated. Virology 2018, 517, 62–68, doi:10.1016/j.virol.2017.11.020.

34. Xu, Y.Z., Thuraisingam, T., Morais, D.A.d.L., Rola-Pleszczynski, M., Radzioch, D. Nuclear translocation of beta-actin is involved in transcriptional regulation during macrophage differentiation of HL-60 cells. Mol. Biol. Cell 2010, 21, 811–820, doi:10.1091/mbc.e09-06-0534.

35. Zhang, X., Goncalves, R., Mosser, D.M. The isolation and characterization of murine macrophages. Curr. Protoc. Immunol. 2008, Chapter 14, Unit 14.1, doi:10.1002/0471142735.im1401s83.

36. Große, M., Ruetalo, N., Businger, R., Rheber, S., Setz, C., Rauch, P., Auth, J., Brysch, E., Schindler, M., Schubert, U. Evidence That Quinine Exhibits Antiviral Activity against SARS-CoV-2 Infection In Vitro. Preprints 2020.

