## Supplementary Figure 1 for "Immune modulating drug MP1032 with SARS-CoV-2 antiviral activity in vitro: A potential multi-target approach for prevention and early intervention treatment of COVID-19"

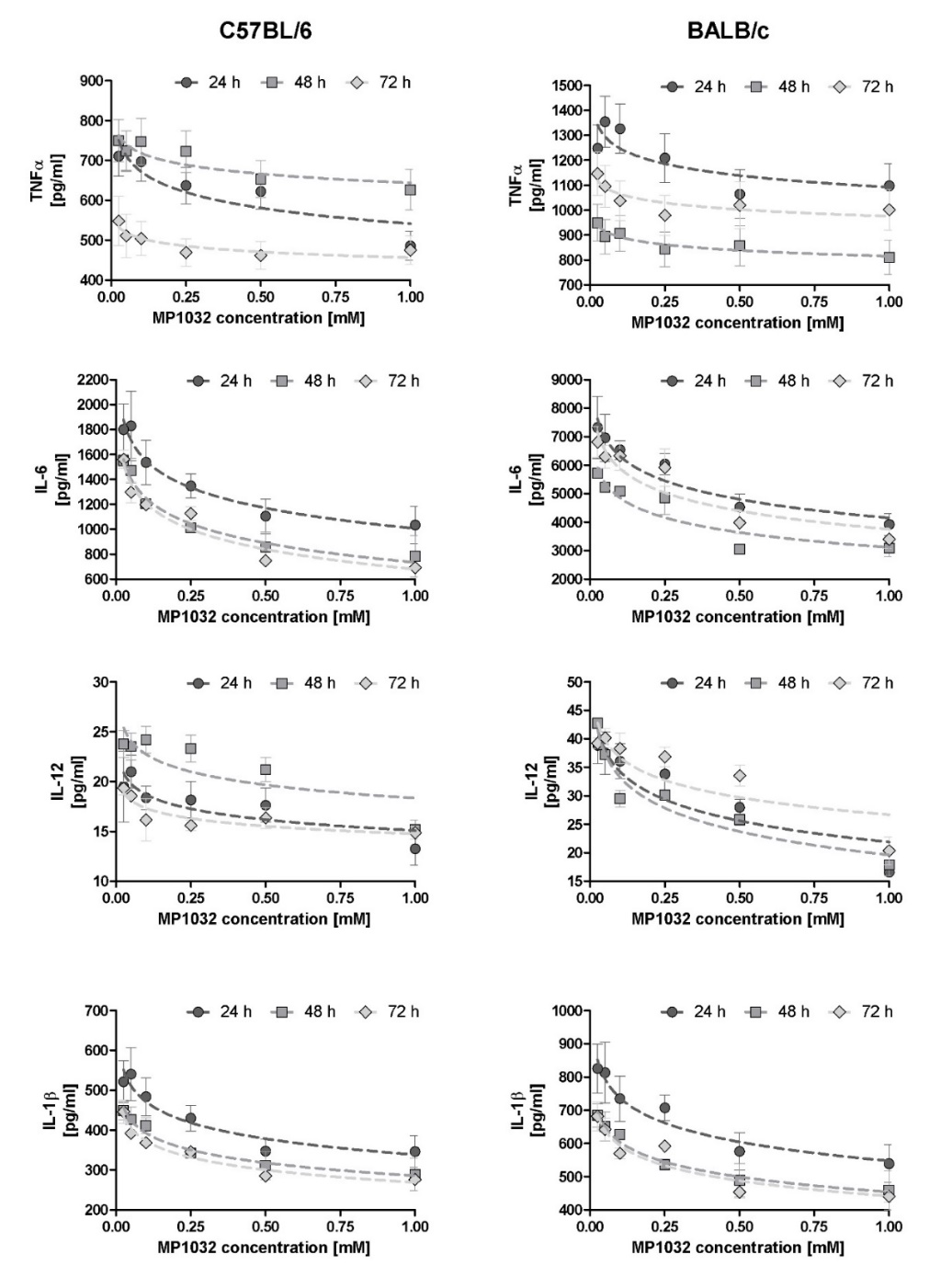

**Supplementary Figure 1: Dose-dependency of MP1032 treatment in murine peritoneal macrophages after 24, 48 or 72 h**

Murine peritoneal macrophages were isolated from female C57Bl/6 and BALB/c mice four days after i.p. injection of 3% thioglycolate medium. One hour prior to LPS stimulation (0.1  µg/ml), isolated cells were pre-treated with different MP1032 concentrations (ranging from 1 mM to 0.025 mM). Cell-free supernatants were collected 24, 48, and 72 h after LPS stimulation and secreted cytokines were detected by ELISA. Data are expressed as mean ±SEM; n=2.
