## Supplementary Table 1 for "Immune modulating drug MP1032 with SARS-CoV-2 antiviral activity in vitro: A potential multi-target approach for prevention and early intervention treatment of COVID-19"

**Supplementary Table 1: Tabulated List of Adverse Drug Reactions.**

Frequency classifications: very common (≥1/10), common (≥1/100 to <1/10), uncommon (≥1/1,000 to <1/100), rare (≥1/10,000 to <1/1,000), very rare (<1/10,000). Coded with MedDRA version 23.0.

| **System Organ Class (SOC)** | **Adverse Reaction (PT – Preferred Term)** | **Seriousness** | **Severity** | **Frequency** |
| --- | --- | --- | --- | --- |
| Blood and lymphatic system disorders | neutropenia | not serious | moderate | uncommon (1/146) |
| Cardiac disorders | palpitations | not serious | moderate | uncommon (1/146) |
| Gastrointestinal disorders | abdominal pain upper | not serious | mild | uncommon (1/146) |
| General disorders and administration site conditions | fatigue | not serious | mild | uncommon (1/146) |
|  | feeling drunk | not serious | mild | uncommon (1/146) |
|  | influenza-like illness | not serious | moderate | uncommon (1/146) |
| Infections and infestations | cystitis | not serious | moderate | uncommon (1/146) |
|  | nasopharyngitis | not serious | moderate | common (2/146) |
| Nervous system disorders | headache | not serious | mild | uncommon (1/146) |
| Skin and subcutaneous tissue disorders | pruritus | not serious | mild to moderate | common (2/146) |
|  | psoriasis | not serious | mild to severe | common (2/146) |
